# Prediction of Procedural Pain during Endometrial Biopsy

**DOI:** 10.1101/2021.03.24.21254143

**Authors:** Hye Gyeong Jeong, Jooyoung Kim, Sung il Choo, Kidong Kim, Banghyun Lee, Soyeon Ahn

**Author notes:** These authors contributed equally to this study (Hye Gyeong Jeong, Jooyoung Kim). Correspondence to: Kidong Kim, MD, PhD, Department of Obstetrics and Gynecology, Seoul National University Bundang Hospital, Seongnam-si, Gyeonggi-do, Republic of Korea.

## Abstract

**Background/Objective:** Endometrial (EM) biopsy is a commonly-performed gynecological procedure that is associated with side effects such as discomfort and pain. The aim of the current study was to predict procedural pain during EM biopsy.

**Methods:** We retrospectively reviewed the medical records of 100 women who underwent EM biopsy between July 2014 and November 2015 in an outpatient clinic of our hospital. Eighty-one patients were included in the final analysis after excluding those who lacked pain data and those who were sedated with midazolam. We examined the association of patient and clinician characteristics with procedural pain, and created a prediction model using characteristics via multiple linear regression analysis.

**Results:** Eighty-one women underwent EM biopsy (dilatation and curettage, EM sampling). In univariable analysis, history of EM biopsy, endometrial thickness (EMT) and training year of operator (TY) were significantly associated with procedural pain. The initial multivariable model was fitted with significant predictors in a univariable analysis. The p-value of EMT and TY was below the pre-defined threshold (0.2) and the final pain prediction model included EMT and TY. Furthermore, pain during the procedure was calculated by the following equation: pain score (numeric rating scale) = 7.364 + (−0.872) * EM thickness (cm) + (−1.033)*TY.

**Conclusion:** Both endometrial thickness and training year of operator were useful predictors of the severity of EM biopsy-related pain.

## Introduction

Endometrial (EM) biopsy is a commonly performed outpatient procedure to evaluate the endometrium in patients with abnormal uterine bleeding or abnormal findings on sonography. According to a report in 2016, 21,889 EM biopsies were performed in South Korea, of which 21071 (91.5%) were performed in outpatient clinics. (1).

EM biopsy causes discomfort and pain. More than half of EM biopsied patients describe their experiences as “moderately” or “severely” painful (2). Previous studies have demonstrated that procedural pain is influenced by parity, pre-procedural anxiety, menopausal status, history of vaginal delivery, provider experience, use of a tenaculum, and procedure time (3, 4). To reduce procedural pain, paracervical block, intrauterine anesthesia, oral medications such as non-steroidal anti-inflammatory drugs or opioids, and intravenous sedation have been used. Many studies have concluded that these methods are effective for reduction of procedural pain (3, 5, 6).

Nevertheless, these methods to reduce procedural pain were not applicable to all patients due to many reasons. For example, intravenous sedation requires patient monitoring and a trained anesthetist as well as appropriate space for recovery after the procedure, and these requirements increase the cost of EM biopsy. If the degree of procedural pain can be predicted prior to EM biopsy, we can implement more cost-effective methods of pain control. Specifically, we can limit the need for intensive, resource demanding anesthesia and analgesia for women who experience moderate or more severe pain.

This study aimed to build a predictive model for procedural pain during EM biopsy.

## Materials and methods

### Patients

This study was approved by the Institutional review board and the requirement for informed consent was waived (B-1606-349-111). We retrospectively reviewed the medical records of 100 women who visited the outpatient clinic of our institute and underwent EM biopsy between July 2014 and November 2015. Patients without a pain record (n=15) and those sedated with midazolam (n=4) were excluded. Finally, a total of 81 patients were included in this study.

### Variables

Age, menopause, parity, history of vaginal delivery, history of EM biopsy, presence of myoma and adenomyosis, uterus size and position, method of EM biopsy (dilatation and curettage or EM sampling), EM thickness, gender and training year of residents who performed the procedure, application of paracervical block, and information regarding maximal pain during procedure were retrieved from the medical records.

All procedures were performed by second or third year residents. The number of second and third year residents was six and three, respectively. During the procedure, the anterior cervical lip was grasped with the tenaculum and then uterine sound was inserted to the uterine fundus. The cervical os was dilated using a Hegar dilator. EM biopsy was performed using a curette or sampler. Immediately after completing the procedure, the resident who performed the EM biopsy recorded information regarding the maximal pain during the procedure and rated this on a 10-point scale (numeric rating scale, NRS). No anesthesia or analgesia was provided before the procedure except for paracervical block in some patients.

### Analysis

All variables except maximal pain during procedure were converted to dichotomous variables. Maximal pain during procedure was summarized into median and inter-quartile range (IQR). The association of variables with maximal pain during procedure was examined using the Mann-Whitney test, and a p-value of <0.05 was considered significant. Cases with unknown values for a variable were excluded from the univariable analysis for that variable. Variables with p-value of <0.05 in univariable analysis were included into multivariable analysis. Variables included in the final model were chosen using backward selection with a threshold of p-value = 0.2. Internal validation of the model was performed using bootstrap analysis based on 1000 replications. Analysis was performed using R 3.3.0 version.

## Results

### Characteristics

Characteristics of patients are summarized in Table 1. The median age of patients was 54 and the majority of women were menopausal (45/81). Most patients had a history of vaginal delivery but only 22 patients had a history of EM biopsy. Myoma and adenomyosis were present in 35 and 9 patients, respectively. The median uterus size was 7 cm and most patients had an antero-verted uterus. Dilatation and curettage was more frequently performed than EM sampling. The median EM thickness was 0.8 cm. Most of the procedures were performed by female, 2^nd^ year residents and paracervical block was performed in over half of patients. The median NRS of maximal pain during the procedure was 4.

**Table 1.**
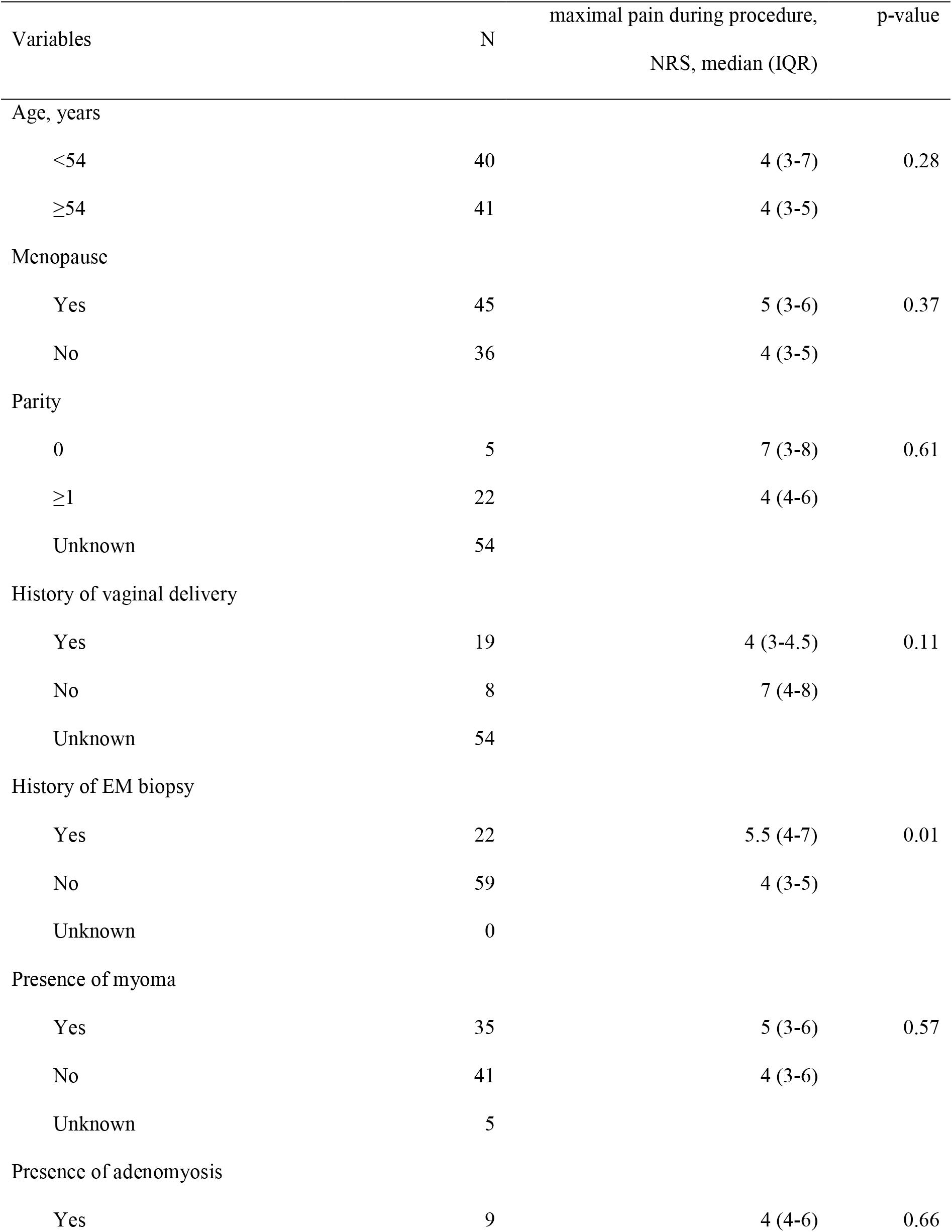
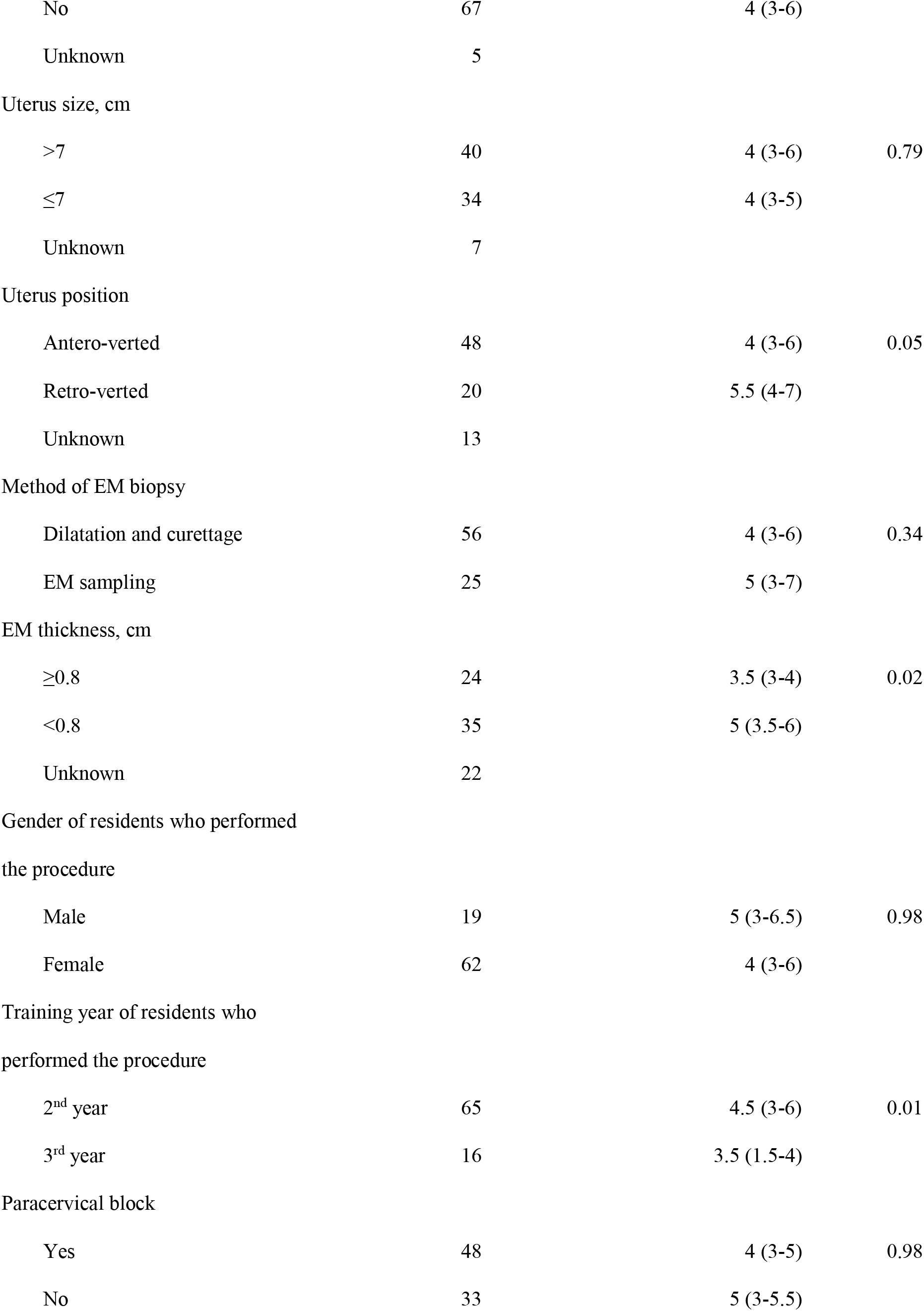
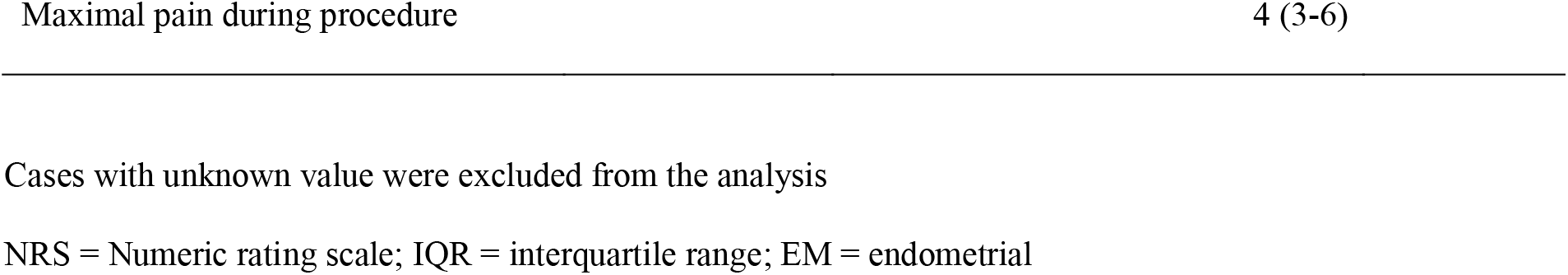
Baseline characteristics and univariable analysis between variables and maximal pain during the procedure (n = 81)

**Table 2.** Multivariable analysis and model development

| Variables in each model | Coefficient | P-value | 95% CI |
| --- | --- | --- | --- |
| Initial model |  |  |  |
| Intercept | 7.092 | <0.01 | 4.214, 9.97 |
| History of EM biopsy | 0.765 | 0.24 | -0.523, 2.052 |
| EM thickness | -0.852 | 0.05 | -1.703, -0.002 |
| Training year of residents who performed the procedure | -0.99 | 0.12 | -2.256, 0.276 |
| Final model |  |  |  |
| Intercept | 7.364 | <0.01 | 4.514, 10.214 |
| EM thickness | -0.872 | 0.05 | -1.725, -0.019 |
| Training year of residents who performed the procedure | -1.0334 | 0.11 | -2.301, 0.235 |

EM = endometrial

### Univariable, multivariable analysis and predictive model

Based on univariable analysis, a history of EM biopsy, EM thickness, and training year of residents who performed the procedure were associated with maximal pain during the procedure (Table 1). Therefore, the initial model was constructed using three variables (history of EM biopsy, EM thickness, and training year of residents who performed the procedure). Because the p-value of history of EM biopsy was over the pre-defined threshold, the final model was constructed using EM thickness and training year of residents who performed the procedure (NRS = 7.364 + (−0.872) * EM thickness (cm) + (−1.033) * training year of residents who performed the procedure (2 vs 3)).

In validation using 1000 times bootstrapping, the mean squared error and mean absolute error of the final model (0.14, 0.16) were better than those of the initial model (0.33, 0.35). The calibration plot of the final model is illustrated in Figure 1.

**Figure 1.**
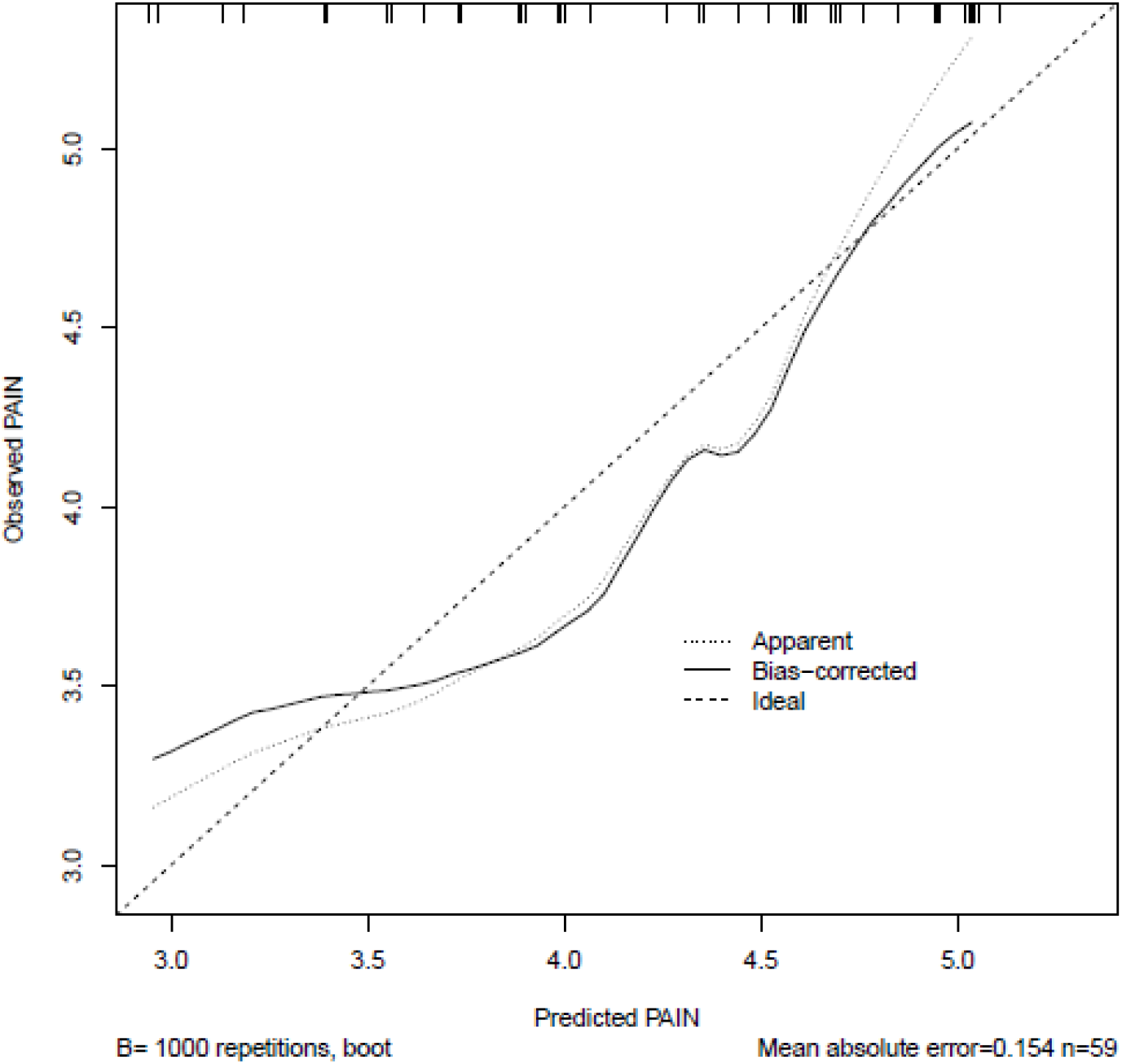
Calibration plot of the final model

## Discussion

We successfully built a predictive model for procedural pain during EM biopsy using EM thickness and training year of residents who performed the procedure. Our model will be useful to help predict procedural pain during EM biopsy and therefore decide whether patients require additional anesthetic or analgesic interventions. According to our model, a thinner endometrium is expected to be associated with a more painful procedure. This suggests that more attention should be given to pain management when an operator performs a biopsy of a thin endometrium. Additionally, our model showed that the level of experience of residents also influences procedural pain, which highlights the importance of appropriately training residents to perform this common procedure.

The results of this study are partially discordant with those of other studies regarding predictors of procedural pain during EM biopsy. For example, in contrast to the results of our study, a previous study reported that postmenopausal women tend to have more severe pain during EM biopsy, and that a history of vaginal delivery was associated with procedural pain (3). A different study also reported a positive correlation between endometrial thickness and pain (<5 mm vs. ≥5 mm) (6), which is contradictory to our results. Consistent with our results, previous studies have demonstrated that the skill or experience of the operator is a predictor of procedural pain (7). In addition in keeping with our results, it has been previously demonstrated that the method of EM biopsy, for example curette versus Pipelle biopsy, was not associated with a significant difference in pain scores (8).

To the best of our knowledge, this is the first study to identify predictors of procedural pain during EM biopsy. Nevertheless, there are also several limitations worth noting. First, this was a single center study and as a result, the number of patients examined was small. Second, this study was vulnerable to many biases because of its retrospective nature. Third, external validation was not performed.

In conclusion, we successfully developed a predictive model for procedural pain during EM biopsy using EM thickness and training year of residents who performed the procedure. We believe this model will help clinicians to manage procedural pain more effectively.

## Data Availability

The minimal data set underlying the findings in our study data is within the paper.

